# Estimates of the COVID-19 pandemic dynamics in Ukraine based on two data sets

**DOI:** 10.1101/2021.02.18.21252000

**Authors:** Igor Nesteruk

## Abstract

**Background:** To simulate how the number of COVID-19 cases increases versus time, various data sets for the number of new cases and different mathematical models can be used. Since there are some differences in statistical data, the results of simulations can be different. Complex mathematical models contain many unknown parameters, the values of which must be determined using a limited number of observations of the disease over time. Even long-term monitoring of the epidemic may not provide reliable estimates of its parameters due to the constant change of testing conditions, isolation of infected and quarantine. Therefore, simpler approaches are necessary. In particular, previous simulations of the COVID-19 epidemic dynamics in Ukraine were based on smoothing of the dependence of the number of cases on time and the generalized SIR (susceptible-infected-removed) model. These approaches allowed to detect the waves of pandemic and to make adequate predictions of the their duration and final sizes. In particular, eight waves of the COVID-19 pandemic in Ukraine were investigated.

**Objective:** We will compare the results simulation of a new epidemic wave in Ukraine based on national statistics and data reported by Johns Hopkins University (JHU).

**Methods:** In this study we use the smoothing method for the dependences of the number of cases on time, the generalized SIR model for the dynamics of any epidemic wave, the exact solution of the linear differential equations, and statistical approach developed before.

**Results:** Ninth epidemic wave in Ukraine was simulated. The optimal values of the SIR model parameters were calculated and compared with the use of two data sets. Both predictions are not very optimistic: new cases will not stop appearing until June-July 2021.

**Conclusions:** New waves of COVID-19 pandemic can be detected, calculated and predicted with the use of rather simple mathematical models. The results of calculations depend on the data sets for the number of confirmed cases. The expected long duration of the pandemic forces us to be careful and in solidarity. The government and all Ukrainians must strictly adhere to quarantine measures in order to avoid fatal consequences. Probably the presented results could be useful in order to estimate the efficiency of future vaccinations.

## Introduction

The studies of the COVID-19 pandemic dynamics in Ukraine are presented in [1-15] and summarized in the book [16]. Different simulation and comparison methods were based on official accumulated number of laboratory confirmed cases [17, 18] (national statistics). These figures coincides with the official WHO data sets, [19]. Unfortunately, WHO stopped to provide the daily information in August 2020.

The classical SIR model [20-22], connecting the number of susceptible *S*, infected and spreading the infection *I* and removed *R* persons, was applied in [2-5, 8-10] to simulate the first pandemic wave in Ukraine. The unknown parameters of this model were be estimated with the use of the cumulative number of cases *V=I+R* and the statistics-based method of parameter identification developed in [23, 24].

The weakening of quarantine restrictions, changes in the social behavior and probably in the coronavirus activity causes change in SIR characteristics and the epidemic dynamics. To detect these changes, a simple method of numerical differentiations of accumulated number of cases was proposed in [11, 15, 16]. To simulate these new pandemic waves, the SIR model was generalized in [12, 15, 16]. In [12, 14, 16] the results of simulation of the first six epidemic waves in Ukraine are presented with the use of a procedure for sequentially determining the parameters of the model for each epidemic wave, starting with the first one.

This method requires considerable effort and time. The book [16] introduced a new algorithm for determining the optimal parameter values for a particular epidemic wave without calculating the dynamics of previous waves and presented calculations for the seventh epidemic wave in Ukraine. The eighth epidemic wave in Ukraine was simulated in [15] with the use of this approach. In this paper, we will analyze the dynamics of the epidemic in Ukraine in the period from September 1 to December 20, 2020, calculate the parameters of the eighth wave and make some predictions.

Since the national statistics does not look complete (see, e.g., the results of total staff testing in two schools and two children gardens in the Ukrainian city of Chelnytskii, [25]), it is very interesting to compare different data sets and simulations results based on different statistical information. For this purpose we will use the data reported by Johns Hopkins University (JHU) [26]. The results of smoothing two data sets and SIR simulations will be presented.

## Materials and Methods

### Data

We will use two data sets regarding the accumulated numbers of confirmed COVID-19 cases in Ukraine: the official information from national sources [17,18] - *V*_*j1*_ and data set *V*_*j2*_ from the COVID-19 Data Repository by the Center for Systems Science and Engineering (CSSE) at Johns Hopkins University (JHU) [26]. These values and corresponding moments of time *t*_*j*_ (measured in days, zero point is November 30, 2020) are shown in Table 1.

**Table 1.**
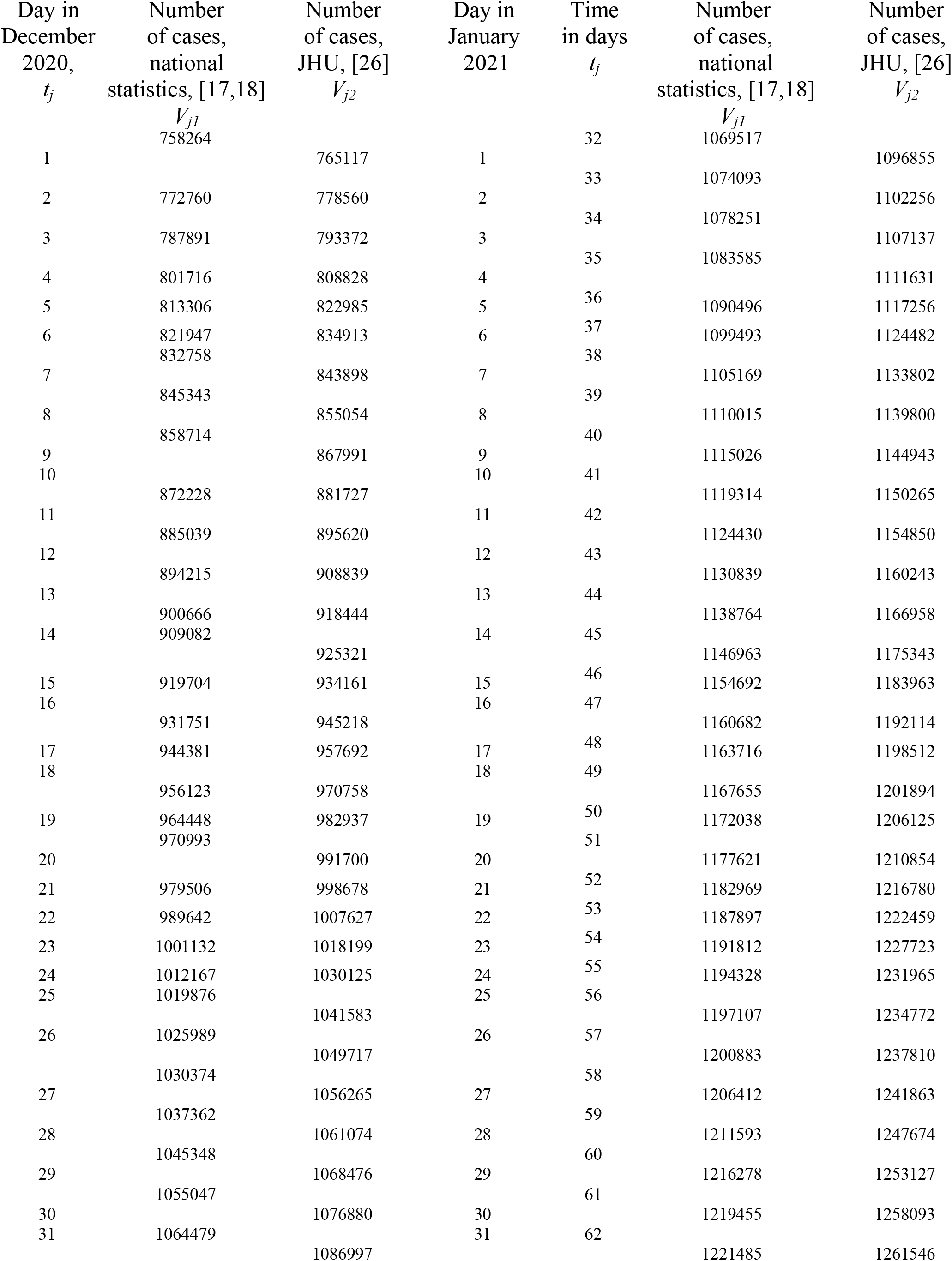
Cumulative numbers of confirmed Covid-19 cases in Ukraine: *V*_*j1*_ -National statistics, [17, 18] and *V*_*j2*_ - according to COVID-19 Data Repository by the Center for Systems Science and Engineering (CSSE) at Johns Hopkins University (JHU), [26].

It must be noted that the data presented in Table 1 does not show all the COVID-19 cases in Ukraine. Many infected persons are not identified, since they have no symptoms. For example, employees of two kindergartens and two schools in the Ukrainian city of Chmelnytskii were tested for antibodies to COVID-19, [25]. In total 292 people work in the surveyed institutions. Some of the staff had already fallen ill with COVID-19 or were hospitalized. Therefore, they were tested and registered accordingly. In the remaining tested 241 educators, antibodies were detected in 148, or 61.4%. Many people know that they are ill, since they have similar symptoms as other members of families, but avoid to make tests. Unfortunately, one laboratory confirmed case can correspond to several other cases which are not confirmed and displayed in the official statistics.

There are also significant differences in two data sets visible in Table 1. The numbers of cases reported by JHU in January 2021 are 30,000-40,000 higher than presented by the national statistics [17, 18]. However, both data sets are probably incomplete. Next, we will make a comparative analysis of these data and conduct appropriate SIR simulations.

### Detection of epidemic waves

To control the changes of epidemic parameters, we can use daily numbers of new cases and their derivatives. Since these values are random, we need some smoothing. For example, we can use the smoothed daily number of accumulated cases proposed in [11, 12, 14-16]:

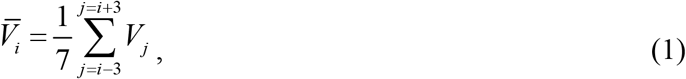

The first and second derivatives can be estimated with the use of following formulas:

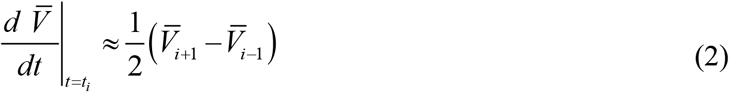

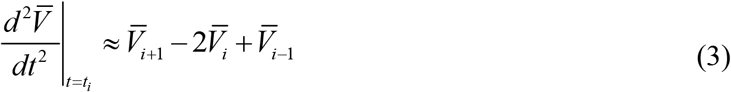

### Generalized SIR model

The classical SIR model for an infectious disease [20-22] was generalized in [12, 14-16] in order to simulate different epidemic waves. We suppose that the SIR model parameters are constant for every epidemic wave, i.e. for the time periods: 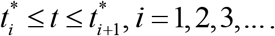. Than for every wave we can use the equations, similar to [20-22]:

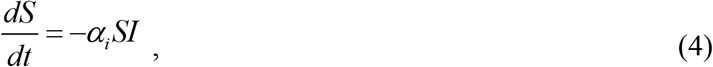

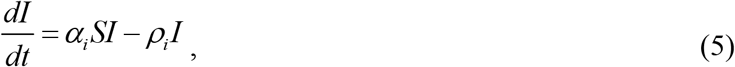

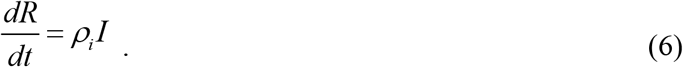

Here *S* is the number of susceptible persons (who are sensitive to the pathogen and **not protected**); *I* is the number of infected persons (who are sick and **spread the infection**; please don’t confuse with the number of still ill persons, so known active cases) and *R* is the number of removed persons (who **no longer spread the infection**; this number is the sum of isolated, recovered, dead, and infected people who left the region). Parameters *α*_*i*_ and *ρ*_*i*_ are supposed to be constant for every epidemic wave.

Parameters *α*_*i*_ show how quick the susceptible persons become infected (see (4)). Large values of this parameter correspond to severe epidemics with many victims. These parameters accumulate many characteristics. First they shows how strong (virulent) is the pathogen and what is the way of its spreading. Parameters *α*_*i*_ accumulate also the frequency of contacts and the way of contacting. In order to decrease the values of *α*_*i*_, we have to minimize the number of our contacts and change our contacting habits. For example, we have to avoid the public places and use masks there, minimize or cancel traveling. We have to change our contact habits: to avoid handshakes and kisses. First, all these simple things are very useful to protect yourself. In addition, if most people follow these recommendations, we have chance to diminish the values of parameters *α*_*i*_effects of the pandemic. and reduce the negative effects of the pandemic.

The parameters *ρ*_*i*_ characterize the patient removal rates, since eq. (6) demonstrates the increase rate of *R*. The inverse values 1/ *ρ*_*i*_ are the estimations for time of spreading infection *τ*_*i*_ during *i-th* epidemic wave. So, we are interested in increasing the values of parameters *ρ*_*i*_ and decreasing 1/ *ρ*_*i*_. People and public authorities should work on this and organize immediate isolation of suspicious cases.

Since the derivative the sum *d* (*S* + *I* + *R*) / *dt* is equal to zero (it follows from summarizing Eqs. (4)-(6)),the sum

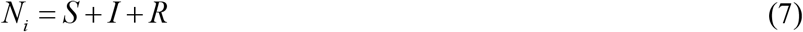

must be constant for every wave and is not the volume of population.

To determine the initial conditions for the set of equations (4)–(6), let us suppose that at the beginning of every epidemic wave 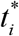 :

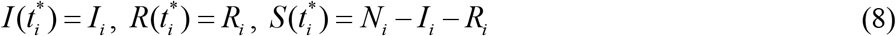

It follows from (4) and (5) that

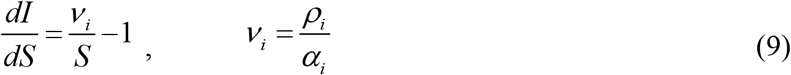

Integration of (9) with the initial conditions (8) yields:

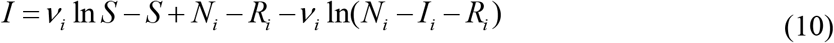

It follows from (9) that function *I* has a maximum at *S* =*ν*_*i*_ and tends to zero at infinity. The corresponding number of susceptible persons at infinity *S*_*i*∞_ > 0 can be calculated from the non-linear equation:

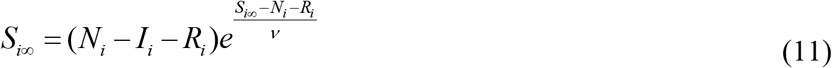

Formula (11) follows from (10) at *I*=0.

In [12, 15, 16] the set of differential equations (4)-(7) was solved by introducing the function

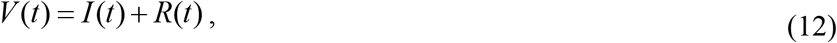

corresponding to the number of victims or the cumulative confirmed number of cases. For many epidemics (including the COVID-9 pandemic) we cannot observe dependencies *S*(*t*), *I* (*t*) and *R*(*t*) but observations of the accumulated number of cases *V*_*j*_ corresponding to the moments of time *t*_*j*_ provide information for direct assessments of the dependence *V* (*t*).

It follows from (5) and (6) that:

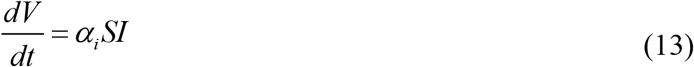

Eqs. (7), (10) and (13) yield:

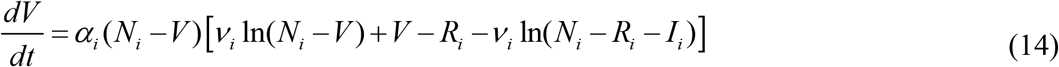

Integration of (14) provides an analytical solution for the set of equations (4)–(6):

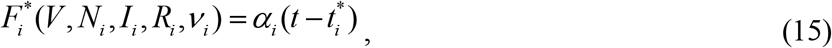

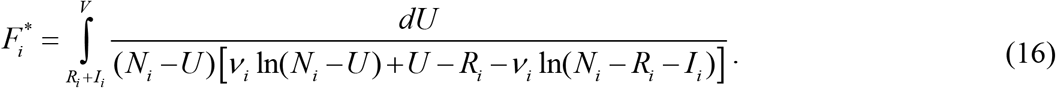

Thus, for every set of parameters *N*_*i*_, *I*_*i*_, *R*_*i*_, *ν*_*i*_, *α*_*i*_ and a fixed value of *V*, integral (16) can be calculated and the corresponding moment of time can be determined from (15). Then functions *I(t)* and *R(t)* can be easily calculated with the use of formulas (10) and:

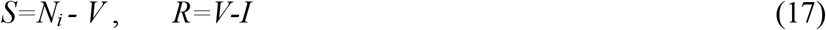

The final numbers of victims (final accumulated number of cases corresponding to the *i-th* epidemic wave) can be calculated from:

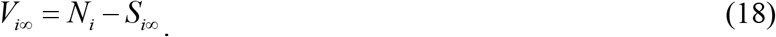

To estimate the final day of the *i-th* epidemic wave, we can use the condition:

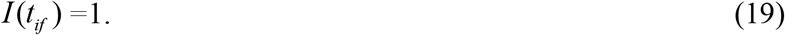

which means that at *t* > *t*_*if*_ less than one person still spreads the infection.

### Parameter identification procedure

In the case of a new epidemic, the values of its parameters are unknown and must be identified with the use of limited data sets. For the first wave of an epidemic starting with one infected person, the number of unknown parameters is only four, since *I*_1_ = 1 and *R*_1_ = 0. The corresponding statistical approach was proposed in [23, 24] and used in [2-5, 8-10] to simulate the first COVID-19 pandemic wave in Ukraine.

For the next epidemic waves (*i* > 1), the moments of time 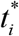 corresponding to their beginning are known. Therefore the exact solution (15)-(16) depend only on five parameters - *N*_*i*_, *I*_*i*_, *R*_*i*_, *ν*_*i*_, *α*_*i*_. Then the registered number of victims *V*_*j*_ corresponding to the moments of time *t*_*j*_ can be used in eq. (16) in order to calculate 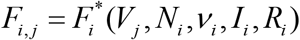 for every fixed values of *N*_*i*_, *ν*_*i*_, *I*_*i*_, *R*_*i*_ and then to check how the registered points fit the straight line (15).

Eq. (15) can be rewritten as follows:

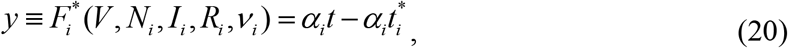

Assuming

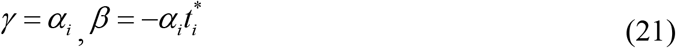

we can estimate the values of parameters *γ* and *β*, by treating the values 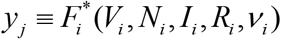 and corresponding time moments *t*_*j*_ as random variables Then we can use the observations of the accumulated number of cases and the linear regression in order to calculate the coefficients 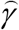 and 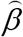 of the regression line

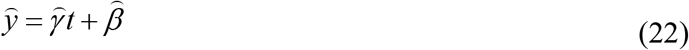

using the standard formulas from, e.g., [27]. Values 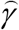 and 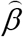 can be treated as statistics-based estimations of parameters *γ* and *β* from relationships (21).

The reliability of the method can be checked by calculating the correlation coefficients *r*_*i*_ (see e.g., [27]) for every epidemic wave checking how close its value is to unity. We can use also the F-test for the null hypothesis that says that the proposed linear relationship (20) fits the data set. The experimental values of the Fisher function can be calculated for every epidemic wave with the use of the formula:

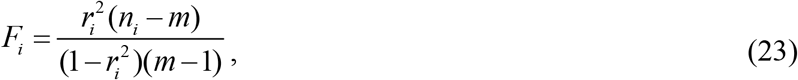

where *n*_*i*_ is the number of observations for the *i*-th epidemic wave, *m* = 2 is the number of parameters in the regression equation. The corresponding experimental value *F*_*i*_ has to be compared with the critical value *F*_*C*_ (*k*_1_, *k*_2_) of the Fisher function at a desired significance or confidence level (*k*_1_ = *m* −1, *k*_2_ = *n*_*i*_ − *m*). When the values *n*_*i*_ and *m* are fixed, the maximum of the Fisher function coincides with the maximum of the correlation coefficient. Therefore, to find the optimal values of parameters *N*_*i*_, *ν*_*i*_, *I*_*i*_, *R*_*i*_, we have to find the maximum of the correlation coefficient for the linear dependence (20). To compare the reliability of different predictions (with different values of *n*_*i*_) it is useful to use the ratio *F*_*i*_ / *F*_*C*_ (1, *n*_*i*_ − 2) at fixed significance level. We will use the level 0.001; corresponding values of *F*_*C*_ (1, *n*_*i*_ − 2) can be taken from [28]. The most reliable prediction yields the highest *F*_*i*_ / *F*_*C*_ (1, *n*_*i*_ − 2) ratio.

The exact solution (15)-(16) allows avoiding numerical solutions of differential equations (4)-(6) and significantly reduce the time spent on calculations. In the case of sequential calculation of epidemic waves *i* = 1,2,3 …, it is possible to avoid determining the four optimal unknown parameters *N*_*i*_, *ν*_*i*_, *I*_*i*_, *R*_*i*_, thereby reducing the amount of calculations and difficulties in isolation a maximum of the correlation coefficient. For parameters *I*_*i*_, *R*_*i*_ it is possible to use the numbers of *I* and *R* calculated for the previous wave of epidemic at the moment of time when the following wave began. Then we need to calculate values 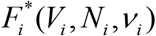, linear regression coefficients (22), correlation coefficient *r*_*i*_, *F*_*i*_ / *F*_*C*_ (1, *n* − 2) and to isolate the values of parameters *N*_*i*_ and *ν*_*i*_ corresponding to the maximum of *r*_*i*_. Knowing the optimal values of five parameters *N*_*i*_, *I*_*i*_, *R*_*i*_, *ν*_*i*_, *α*_*i*_, the SIR curves and other characteristics of the corresponding epidemic wave can be calculated with the use of formulas (10)-(17). This approach has been successfully used in [12, 14-16]. In particular, six waves of the Covid-19 epidemic in Ukraine and four pandemic waves in the world were calculated.

Segmentation of epidemic waves and their sequential SIR simulations need a lot of efforts. To avoid this, a new method of obtaining the optimal values of SIR parameters was proposed in [15, 16]. First of all we can use the relationship

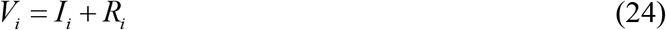

which follows from (12). To estimate the value *V*_*i*_, we can use the smoothed accumulated number of cases (e.g., formula (1)). Then

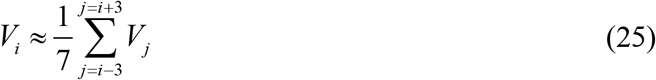

where *i* corresponds to the moment of time 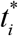. To obtain one more relationship, let us use (7) and (13)

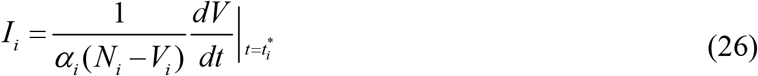

To estimate the average number of new cases *dV/dt* at the moment of time 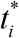, we can use (2). Thus we have only two independent parameters *N*_*i*_ and *ν*_*i*_. To calculate the value of parameter *α*_*i*_, some iterations can be used (see details in [15, 16]).

## Results and discussion

The COVID-19 pandemic characteristics for Ukraine in December 2020 and January 2021 are shown in Figs.1 and 2. Red color corresponds to the national statistics [17,18]; black - to JHU data [26]. “Circles” show the corresponding accumulated numbers of cases; lines represent the smoothed number of accumulated cases (eq. (1)); “crosses” – the first derivative (eq. (2)) multiplied by10; “dots” – second derivative (eq. (3)) multiplied by 1000. Fig. 1 illustrate the data sets presented in Table 1. It can be seen one day shift for the values of the second derivatives calculated with the use of different data sets. If the numbers of cases reported by JHU are attributed to the previous day, the differences in the values of the second derivatives become almost imperceptible (see Fig. 2), but the numbers of accumulated cases according to JHU are still much higher than for the national statistics (see Fig. 2).

**Fig 1.**
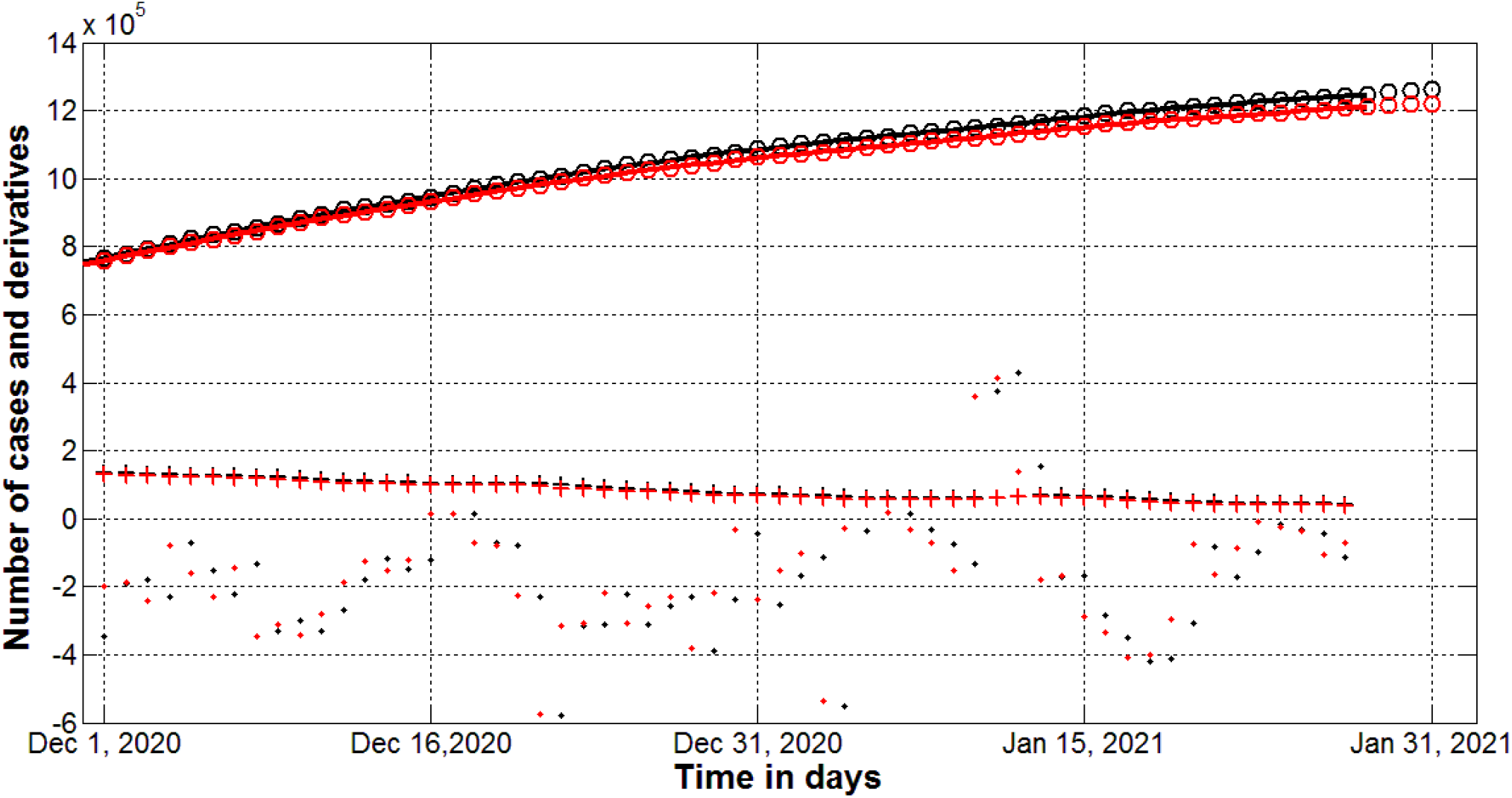
Pandemic dynamics in Ukraine in December 2020 and January 2021 calculated with the use of data sets from Table 1: national statistics [17, 18] (red) and data set reported by JHU [26] (black). Accumulated number of cases - “circles”; smoothed values - lines, eq. (1)). “Crosses” show the first derivatives (eq. (2)) multiplied by 10, “dots” - the second derivative (eq. (3)) multiplied by 1000.

**Fig 2.**
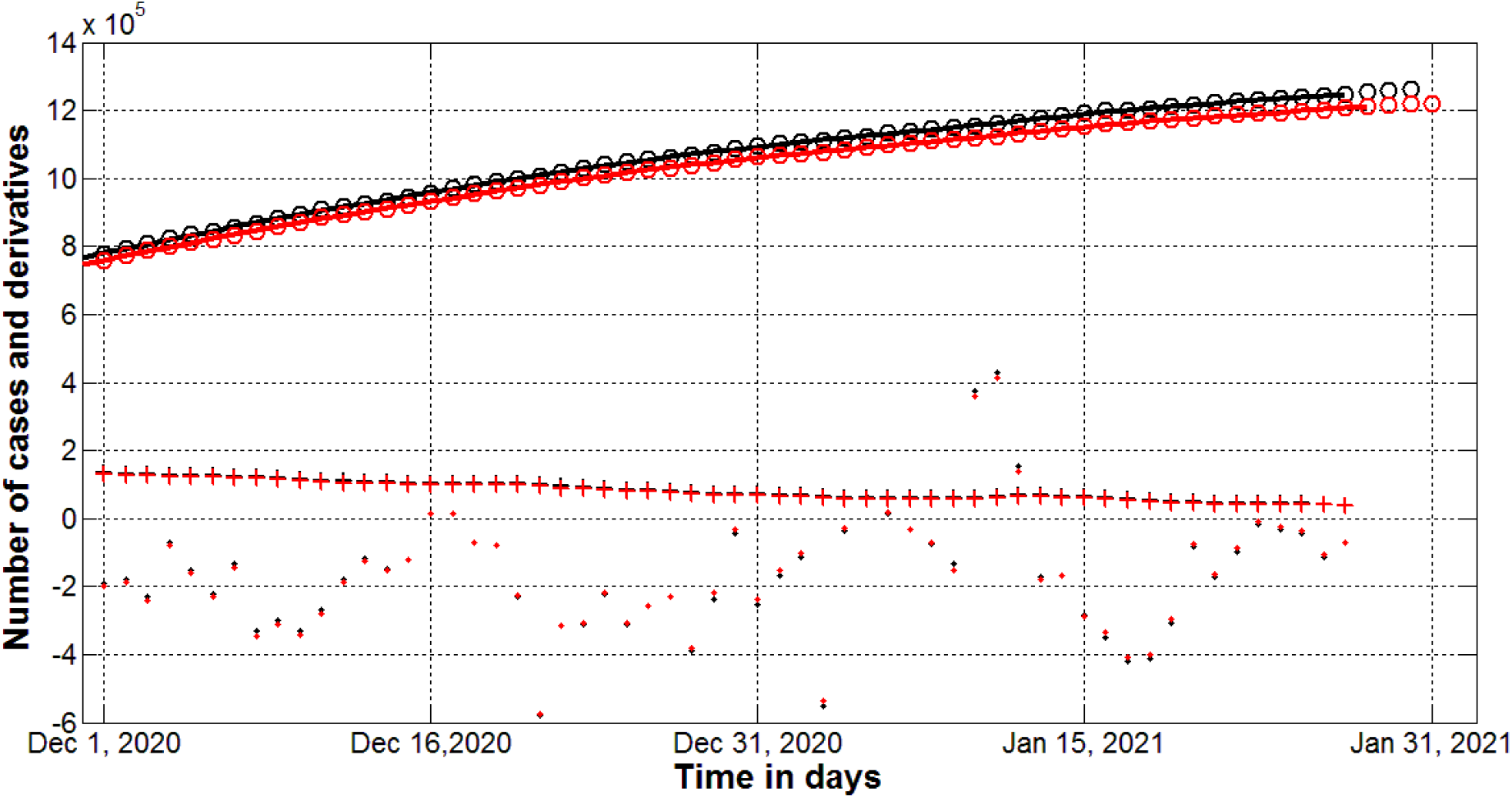
Pandemic dynamics in Ukraine in December 2020 and January 2021 with the numbers of cases reported by JHU attributed to the previous day: national statistics [17, 18] (red) and data set reported by JHU [26] (black). Accumulated number of cases - “circles”; smoothed values - lines, eq. (1)). “Crosses” show the first derivatives (eq. (2)) multiplied by 10, “dots” - the second derivative (eq. (3)) multiplied by 1000.

The jump of the second derivatives occurred on January 10-12, 2021 (see “dots” in Fig. 2) can be explained the New Year and Christmas celebrations. The increase in the number of contacts caused the increase in the number of new cases after some incubation period (see “crosses” in Figs. 1 and 2). The national lockdown in the period January 8-24 allowed stopping this tendency.

The results of SIR simulations with the use of two data sets presented in Table 1 are shown in Table 2 and Fig. 3. Since eight epidemic wave was already calculated in [12, 14-16], we took the period December 11-24, 2020 to calculate the ninth wave with the use of two datasets (presented in Table 1). The optimal parameters of SIR model and other epidemic characteristics can be compared with the results obtained in [15] for the eighth wave (see Table 2). The number of observations taken for calculations *n*_*i*_ was 14 in all cases.

**Table 2.**
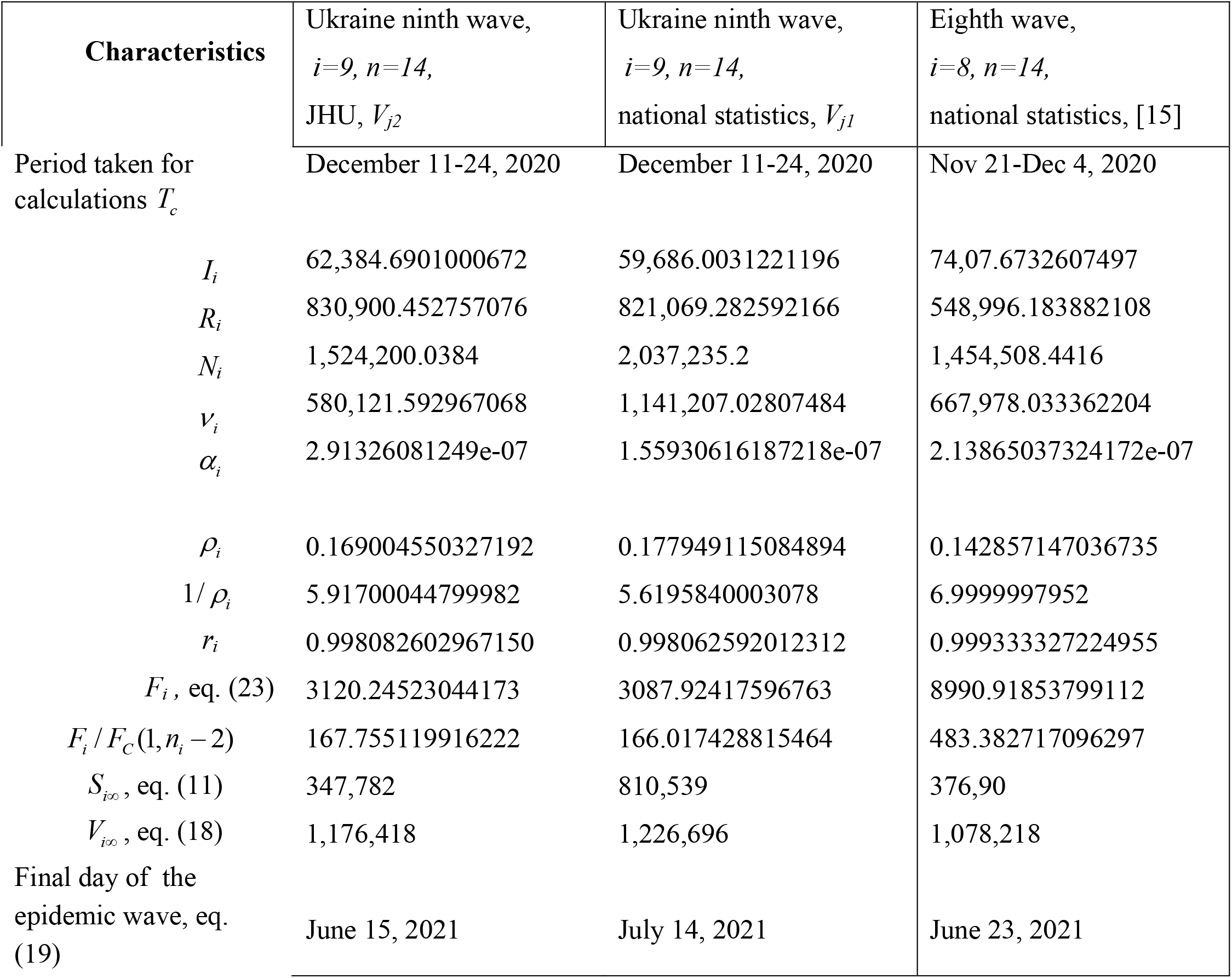
Calculated optimal values of SIR parameters for the eighth and ninth waves of the COVID-19 epidemic in Ukraine.

**Fig 3.**
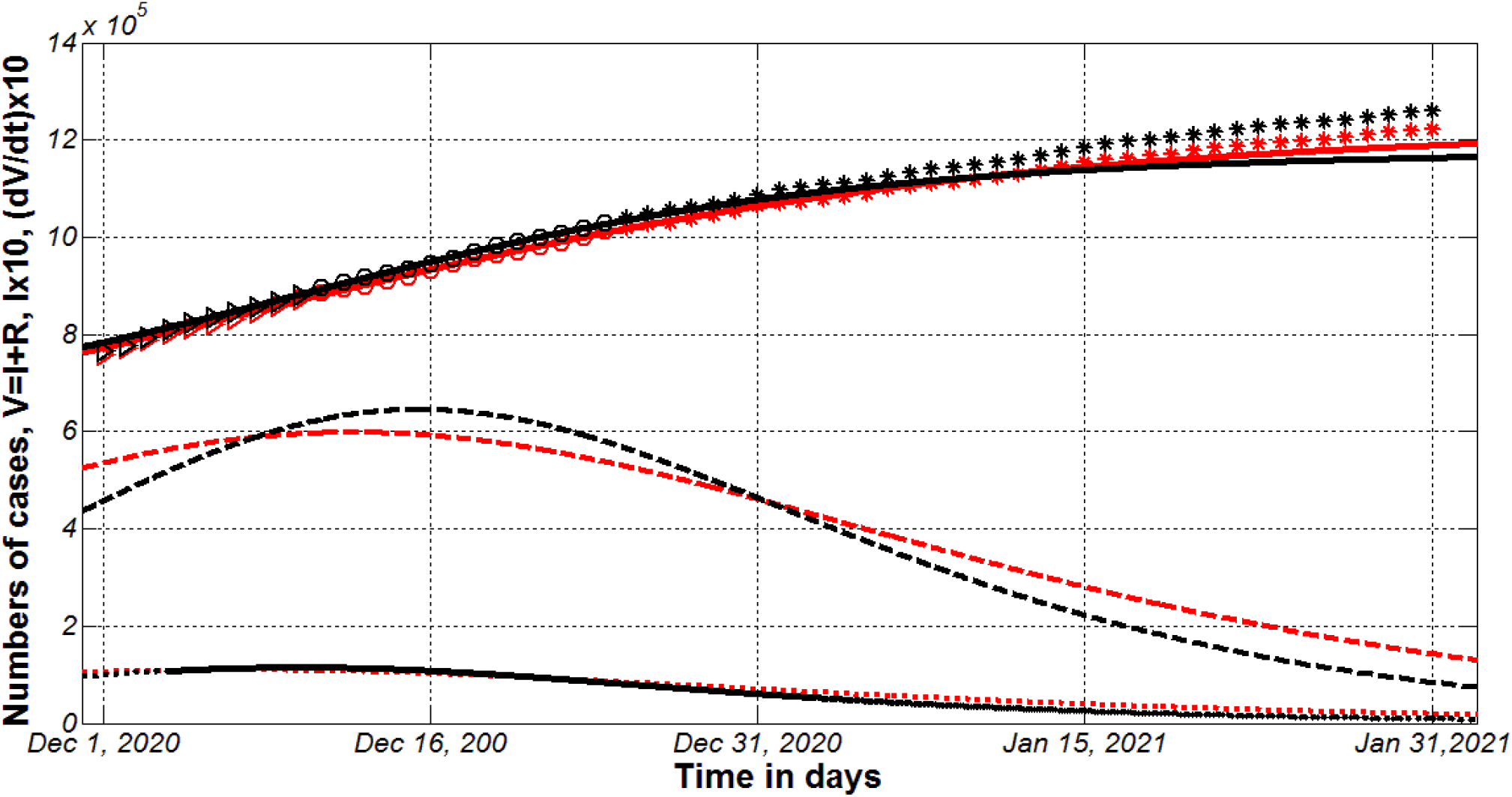
The ninth COVID-19 epidemic wave in Ukraine. SIR simulations (lines) with the use with the use of data sets from Table 1: national statistics [17, 18] (red) and data set reported by JHU [26] (black). Numbers of victims *V(t)=I(t)+R(t)* – solid lines; numbers of infected and spreading *I(t)* multiplied by 10 – dashed; derivatives *dV/dt* (eq. (13)) multiplied by 10 – dotted. Markers show accumulated numbers of cases *V*_*j1*_ and *V*_*j2*_ from Table 1. “Circles” correspond to the accumulated numbers of cases taken for calculations (during period of time *T*_*c*_); “triangles” – numbers of cases before *T*_*c*_; “stars” – number of cases after *T*_*c*_.

It can be seen two data sets yield rather different values of the optimal parameters for the ninth wave (especially for *N*_*9*_, *S*_9∞_, and *ν*_9_), nevertheless the final sizes of this wave *V*_9∞_ and *ρ*_9_ are rather close; the duration based on the national statistics is one month longer in comparison with the calculations based on JHU data set. Both simulations for the ninth epidemic wave in Ukraine yield slightly higher final sizes in comparison with the eighth wave (see Table 2).

Fig. 3 illustrates the results of SIR simulations of the ninth epidemic wave in Ukraine obtained with the use of two data sets: national statistics [17, 18] (red) and data set reported by JHU [26] (black). Numbers of victims *V(t)=I(t)+R(t)* (eq. (12)) are represented by solid lines. Numbers of infected and spreading the infection persons *I(t)* are shown by dashed lines. Dotted lines represent the derivatives *dV/dt* calculated with the use of eq. (13). This derivative yields the estimation of the daily number of new cases. Markers show accumulated numbers of cases *V*_*j1*_ and *V*_*j2*_ from Table 1. “Circles” correspond to the accumulated numbers of cases taken for calculations (during period of time *T*_*c*_); “triangles” – numbers of cases before *T*_*c*_; “stars” – number of cases after *T*_*c*_. It can be seen that the accuracy of simulations based on the national statistics is rather good (the deviations between red “stars” and red solid line are small). The use of JHU data sets yields worth accuracy. Nevertheless, the real numbers of cases already exceed the predicted saturations levels *V*_9∞_ for both data sets and corresponding simulations. As of February 14, 2021 the national statistics yield the figure 1,271,143 of accumulated cases in Ukraine [17, 18]. Thus the epidemic observations during 52 days (after *T*_*c*_) demonstrated only 3.5% exceeding of the saturation level in the case of national statistics.

## Conclusions

Constant changes in the Covid-19 pandemic conditions (i.e., in the peculiarities of quarantine and its violation, in situations with testing and isolation of patients, in coronavirus activity due to its mutations etc.) lead to new pandemic waves and cause the changes in the values of parameters of the mathematical models. Incomplete data and different methods of cases recording can give quite different values of model parameters and forecasts, which was demonstrated by the example of two data sets for Ukraine. The forecast of the duration of the epidemic in Ukraine is not optimistic. New cases will appear at least until July 2021. Let us hope that the number of new cases will grow slowly according to the results of both SIR simulations. To maintain the existing positive dynamics, it is necessary to adhere to quarantine measures and social distancing. Pandemic surveillance experience shows that neglecting simple precautions immediately leads to new waves.

The obtained results can be also useful for assessing the effectiveness of mass vaccination in Ukraine and other countries.

## Data Availability

All data is in text

## Acknowledgements

I am very grateful to Professor Noureddine Benlagha and Oleksii Rodionov for their support and help in collecting and processing data.

## Notes

### Competing Interest Statement

The authors have declared no competing interest.

### Funding Statement

no funding

